# Listract test: a Standardized Assessment Method for Isolated Lisfranc Instability in Cadaver Models

**DOI:** 10.1101/2023.01.16.23284641

**Authors:** Vasundhara Mathur, David O. Osei-Hwedieh, Sayyed Hadi Sayyed Hosseinian, Fernando Raduan, Philip Kaiser, Hamid Ghaednia, Gregory Waryasz, Lorena Bejarano-Pineda, John Kwon, Soheil Ashkani-Esfahani

## Abstract

**Background:** The gold standard for diagnosis of Lisfranc instability is direct visualization in the operation room while the examination techniques is still unstandardized and non-reproducible. We aimed to introduce a novel reproducible intraoperative mechanical testing method (Listract test) for intraoperative isolated Lisfranc instability assessment.

**Methods:** The Lisfranc ligament between the first cuneiform (C1) and second metatarsus (M2) in eight lower leg cadaveric specimens were dissected to replicate C1-M2 Lisfranc instability by eight foot and ankle surgeons. The 50N distraction force was applied in the direction of the C1-M2 ligament. Three methods of fixation - flexible fixation, metal screw, and bio-absorbable screw- were used to fix the injury, and Listract test was applied again after fixation. Besides intraoperative assessment, C1-M2 diastasis and area were measured using radiographs for assessment of Lisfranc instability.

**Results:** The sensitivity and specificity of the Listract test for detection of C1-M2 instability were 100% and 100% intraoperatively, 33.3% and 95.2% using radiographic diastasis measurement, and 63.2% and 38% using area measurement, respectively. The Listract test had a specificity and sensitivity of 100% and 96% for intraoperative assessment, 87.5% and 64.3 for radiographic C1-M2 diastasis, and 48% and 50% for radiographic area.

**Conclusion:** The Listract test is a simple, reproducible, and replicable intraoperative method for evaluating the Lisfranc joint for instability. Developing a device with this mechanism can help clinicians confirm the diagnosis and provide appropriate treatment particularly for equivocal diagnoses.

**Authors’ contributions:** SAE: Conception, design, and conducting the study, analysis, drafting the manuscript; VM, DOH, SHSH, FR: conducting the study, analysis, preparing the manuscript draft; LBP, JK, GW, PK: conducting the study, manuscript preparation, discussion.

## Introduction

Injuries to the Lisfranc joint represent 0.2% of all fractures and have an annual incidence of 1/55,000 individuals.^1,2^ Ligamentous type of Lisfranc injuries is difficult to detect, with 20-40% of them remaining undetected or misdiagnosed at initial presentation.^3^ In such cases, debilitating sequelae like midfoot instability, arch collapse, and traumatic arthritis are more likely to follow, thereby underlying the importance of timely diagnosis and treatment.^4^ Surgical intervention is key to the preservation of the joint and to achieving anatomic reduction.^5,6^ A high portion of instabilities caused by joint widening is observed between the first cuneiform and second metatarsal bones (C1-M2), and between the first and second cuneiform bones (C1-C2). Various types of metal and flexible fixations are used as fixation methods for Lisfranc instabilities.^7,8^ While magnetic resonance imaging (MRI) and weight-bearing CT (WBCT) have shown high sensitivity and specificity for the detection of Lisfranc instability, radiographs, especially in weight-bearing status, are still the mainstay for primary assessment. Despite various imaging methods, intraoperative examination is the gold standard for confirmation of the diagnosis.^9,10^ However, the intraoperative stress examination of the Lisfranc joint by applying a distraction force is not standardized and not a reproducible method.^11^ Surgeons apply various amounts of force on the joint to confirm the diagnosis. This can lead to over or under-diagnosis of the instability. Aiming to introduce a more standardized and dynamic assessment method, a study suggested using Freer to apply stress on the joint. Authors in this study suggested that when the Freer elevator on the thin side could be twisted >90 degrees on the wide side, the Lisfranc joint could be considered unstable.^12^ It is also assumed that the distracting forces applied to the joint should be in the direction of the forces that the actual ligaments bear in physiologic weight-bearing condition.^13^

In this study we aimed to introduce a standardized and reproducible intraoperative mechanical test for Lisfranc instability, particularly in isolated Lisfranc ligament injuries. Our hypothesis is that resembling a standard distracting force applied on the Lisfranc ligament in the physiologic weight-bearing position can result in a reproducible and more accurate evaluation method.

## Materials and Methods

All experiments in this study were approved by the Institutional Research Board protocol (IRB #2016P001295). Fresh-frozen lower-leg cadaveric specimens amputated from the proximal tibia were completely thawed passively prior to conducting the experiment. Radiographic images were obtained in various stages including before injury (stable Lisfranc joint), after injury (unstable Lisfranc after C1-M2 ligament dissection), and after each fixation (Stabilized Lisfranc joint) with and without Listract test. The C1-M2 diastasis and area were measured on these images at each stage. Figure 1 shows the stepwise process of the experiment. For induction of the instability, a complete dissection of the C1-M2 ligament was made from the dorsal to the plantar surface of the foot.

**Figure 1.**
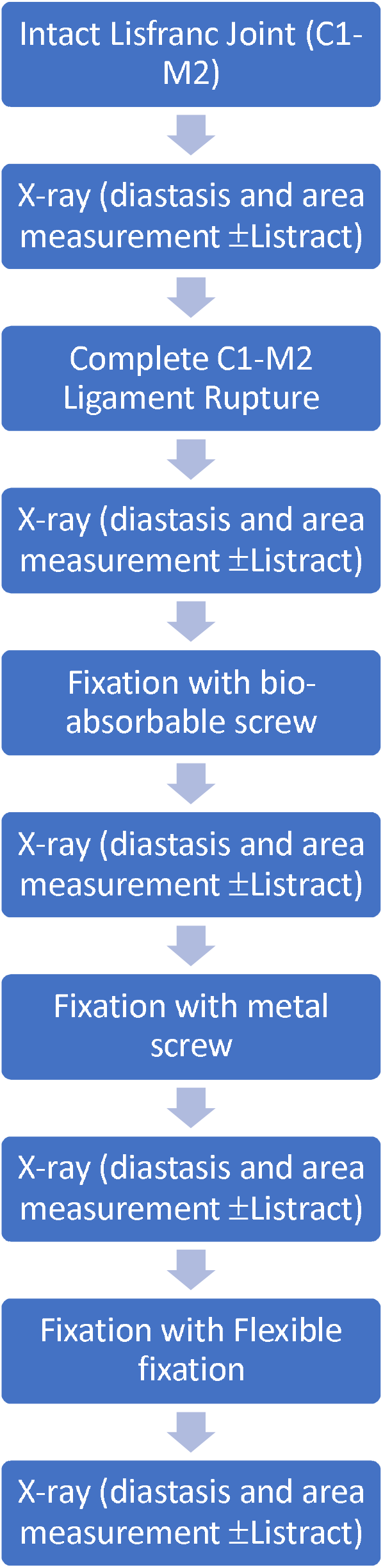
The stepwise process of the experiment assessing the use of the Lisfranc distraction test (Listract test) for detection of isolated Lisfranc ligament rupture leading to instability.

To apply the “Listract test” in the direction of the C1-M2 ligament, a K-wire was drilled through C1 from the dorsal to the plantar aspects of the foot at a 90° angle to the bone. Another K-wire was drilled through the base of M2 dorsal to plantar at a 90° angle to the bone. The positions of the K-wires were confirmed via radiographic images. These K-wires served as opposite pivot points to apply the distraction force between C1 and M2. Using a radiolucent wire that could tolerate a force of ∼15 kg (33.1 lbs), the K-wires were pulled with a 50N (5 kg or 11.02 lbs). Figure 2 shows the construction of the test using K-wires, radiolucent wires (fish wire), and pulleys to direct the forces from both opposite directions. To select the amount of force needed to conduct the test, in a pilot study on 4 cadaver specimens prior to the main experiment, we used different distraction forces, including 25N (2.5 kg), 50N (5 kg), and 100N (10 kg). We considered 2 mm C1-M2 diastasis as the threshold for instability. The 25N forces from two directions were not sufficient to render 2 mm diastasis in unstable Lisfranc joints while the 100N force, though led to >2 mm widening in all four specimens, were too heavy, making the test hard to use in practice and to keep the feet steady while applying the forces. Thus, we selected 50N forces for the main experiment since it led to >2 mm widening in all the specimens and was also feasible to apply in practice. We applied the Listract test on the specimens in five stages (Figure 1), including 1) Intact Lisfranc ligament, 2) full resection of Lisfranc ligament leading to instability of the joint, and 3) fixation with bioabsorbable radiolucent screws (OSSIO Inc., MA, USA), 4) fixation with single cortical metal screws (DePuy-Synthes, West Chester, PA), 5) fixation with single flexible fixation method (mini-Tightrope, Arthrex, Naples, FL, US). Radiographic images were obtained in ten stages, including a) intact Lisfranc ligament with and without applying the Listract test, b) dissected Lisfranc C1-M2 ligament ± Listract test, c) bio-absorbable screw fixation ± Listract test, d) metal screw fixation ± Listract test, e) flexible fixation ± Listract test. Eight orthopaedic foot and ankle surgeons performed the experiment on separate cadavers. We could not perform direct measurement of the C1-M2 diastasis due to the presence of K-wires that barred the C1-M2 region. Three independent observers performed measurements on all radiographs. These three observers were unaware of the procedures and differences in the forces applied on the joint in each imaging stage. The measurements included C1-M2 diastasis and C1-M2 area on dorsiplantar radiographic images.

**Figure 2.**
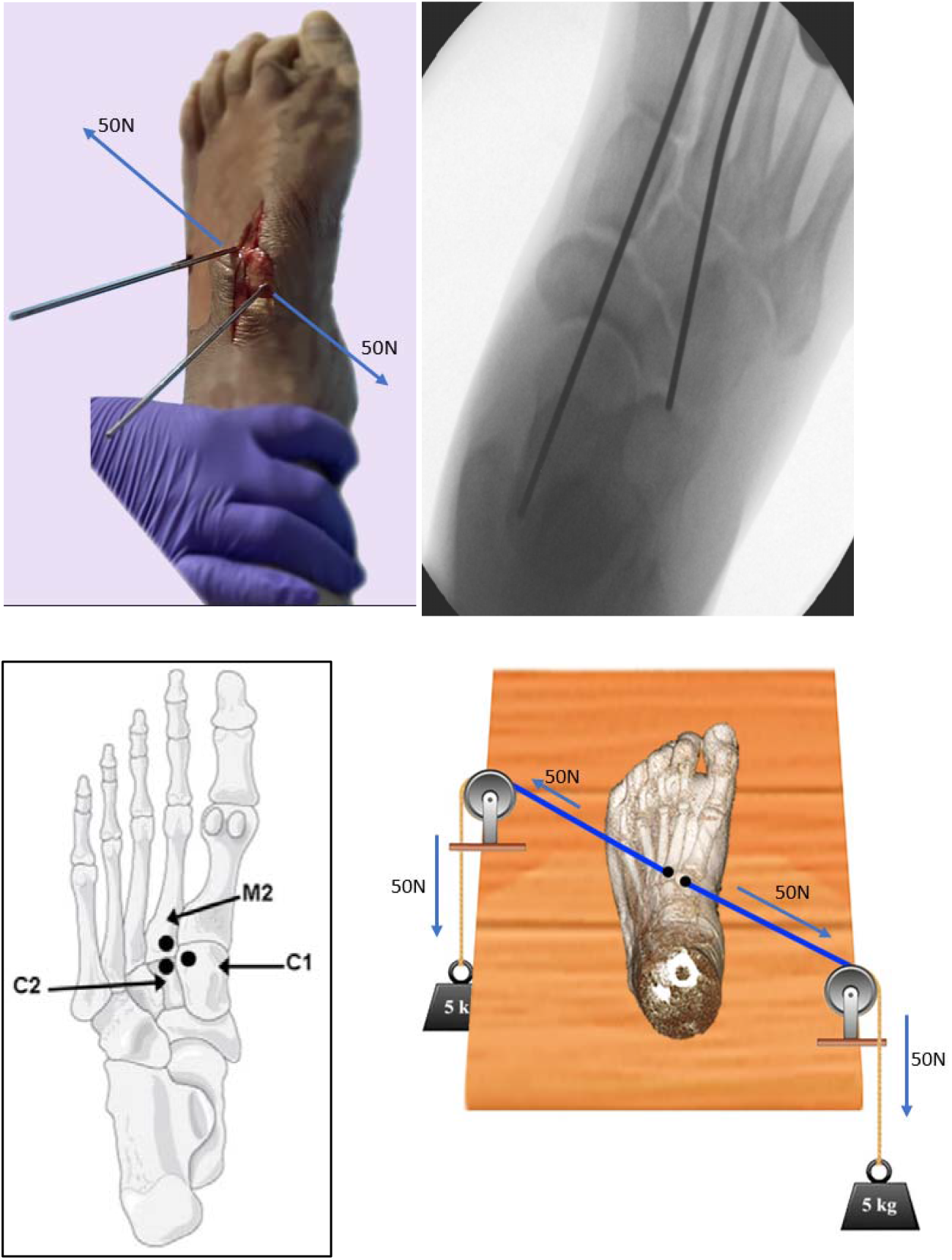
The Listract test; a mechanical distraction test exerted in alignment with the direction of the C1-M2 ligament to assess the stability of the joint. A schematic view of a cadaveric left foot subjected to the distraction forces applied using a pulley system.

### Statistical Analysis and Interobserver Reliability

C1-M2 diastasis and area measurements were performed in each stage by three expert observers and reported as the mean and standard deviation (SD). Comparisons of the measurements in each stage on the radiographic images were performed using Mann-Whitney U test. Comparison of the measurements after the three fixation methods and the measurements performed at the dissected ligament stage were compared using the Kruskal-Wallis test. P-value <0.05 was considered statistically significant. For these analyses, SPSS software (version 26.0, Armonk, NY, USA) was used for data analysis in this study. The three observers were asked to assess the joint instability intraoperatively before and after dissecting the C1-M2 ligament while they were blinded to the condition of the ligament. Their answers were recorded. Additionally, the sensitivity and specificity values for each intraoperative diagnosis and each measurement, diastasis, and area, in detecting Lisfranc instability with and without Listract test were calculated. The cut-off value for diastasis was considered 3mm, and for area was 30mm^2^.^14,15^ For diastasis and area measurements among the observers with and without the Listract test, we used the intraclass correlation coefficient (ICC). Values less than 50%, between 50% and 75%, between 75% and 90%, and greater than 90% were considered poor, moderate, good, and excellent, respectively.

## Results

The mean and SD for all stages of measurements have been tabulated (Table 1). Applying Listract test to the unstable C1-M2, led to a significant difference between area with Listract compared to the area sans Listract test. Applying Listract test to C1-M2 after flexible fixation led to a significant difference in the diastasis before and after the test (Table 1). The measurements between the unstable C1-M2 and the measurements after all fixation methods were significantly different with and without the Listract test (P<0.05).

**Table 1.** Diastases and areas of C1-M2 in cadaver specimens after each stage, including intact and dissected Lisfranc ligament, and after fixation with flexible, metal screw, and bio-absorbable fixations. Each measurement was conducted without and with the Listract test. Data are shown as mean (SD).

| *Stage* | *Diastasis*<br>*(mm)* | *Diastasis +*<br>*Listract test* | *P-value<sup>†</sup>* | *Area(mm<sup>2</sup>)* | *Area + Listract test* | *P-value<sup>†</sup>* |
| --- | --- | --- | --- | --- | --- | --- |
| *Stable C1-M2* | 1.61 (0.27) | 1.82 (0.57) | 0.11 | 26.8 (6.21) | 31.3 (11.06) | **<0.001** |
| *Unstable C1-M2* | 2.47 (0.51) | 2.75 (0.52) | 0.13 | 32.7 (13.08) | 41.8 (14.32) | **0.002** |
| *P-value\** | **<0.001\*** | **<0.001\*** |  | **0.001\*** | **0.001\*** |  |
| *Flexible fixation* | 2.02 (0.50) | 2.22 (0.24) | **0.03** | 30.75 (7.42) | 29.9 (6.8) | 0.09 |
| *P-value* | **0.01\*\*** | **0.04\*\*** |  | **0.03\*\*** | **<0.001\*\*** |  |
| *Metal Screw*<br>*fixation* | 1.72 (0.63) | 1.61 (1.31) | 0.40 | 30.75 (17.13) | 31.8 (19.19) | 0.08 |
| *P-value* | **<0.001\*\*** | **<0.001\*\*** |  | **0.01\*\*** | **0.01\*\*** |  |
| *Bio-absorbable*<br>*screw fixation* | 1.67 (0.77) | 1.69 (0.64) | 0.50 | 29.53 (9.15) | 31.13 (9.75) | 0.06 |
|  | **<0.001\*\*** | **<0.001\*\*** |  | **0.01\*\*** | **0.003\*\*** |  |

<sup>†</sup> Measurement with vs. without Listract test, Mann-Whitney U test; p-value<0.05 considered significant.
\* Measurements in intact vs. dissected Lisfranc ligament; Mann-Whitney U test.
\*\* Measurements in the fixation method vs. dissected Lisfranc ligament stage; Mann-Whitney U test.

The ICC for C1-M2 instability diagnosis intraoperatively with Listract test was 0.97 (95%CI=0.94-0.99). For the C1-M2 diastasis (0.64, 95%CI=0.53-0.73) and area (0.80, 95%CI=0.72-0.86) measurements were moderate and good, respectively. The sensitivity and specificity for intraoperative Listract test were 0.96 and 1.00. The specificity and specificity for radiographic diastasis were 0.15 and 1.00 without Listract and 0.33 and 0.95 with Listract test, respectively. The specificity and specificity for area measurement were 0.63 and 0.38 without Listract and 0.67 and 0.76 with Listract test, respectively.

## Discussion

The purpose of this study was to introduce a standardized, reproducible, and relibale examination method for Lisfranc instability diagnosis. Despite various radiological methods introduced to detect Lisfranc instability, particularly for C1-M2 and C1-C2 instabilities, the intraoperative diagnosis remained the gold-standard method for confirmation. However, the methods used by most of the surgeons were not standardized, not based on a specific amount of force, and thus, not reproducible. Given diastasis and area measurement as two radiographic methods for the detection of C1-M2 instability, we found that Listract test can increase the specificity and sensitivity of these tests using pre-defined cut-off values. Listract test applies a specific amount of force (50N) on the joint in the direction of the ligaments. This helps the surgeon mimic the physiologic load on the joint intraoperatively and assess the joint in a condition closer to real-life conditions. Moreover, Listract test has led to greater values both for diastasis and area; however, only the area showed a significant difference when comparing measurements with versus without Listract measurements. Future studies can also focus on introducing an intraoperative cut-off value for the diastasis to increase.

All the fixation groups showed significant differences with the unstable group and values similar to the stable groups. This can indicate similar efficacy of all the fixation methods for Lisfranc instability including the novel bio-absorbable screws. While the Listract test produced a nearly perfect ICC (97%) and it showed to increase in the sensitivity and specificity of both radiographic diastasis and area measurements, the need to reassess the cut-off values for both of these radiographic measurements seems inevitable. The interobserver agreement was moderate and good for diastasis and area measurement, respectively; however, previous reports using 3D weight-bearing CT scans have shown an ICC of >0.96 using 3D volume measurement for this non-invasive imaging method ^10,16–18^. Bhimani et al. have shown that weight-bearing CT scan can detect Lisfranc instability in C1-M2 joint with a sensitivity and specificity of 0.79 and 96.5 for diastasis and area, respectively.^10^ They have also shown that 3D volume measurement can have a specificity and sensitivity of 0.98 and 0.92, respectively. However, performing weight-bearing CT scan in patients who cannot tolerate the pain or in the operative room is not feasible. Kitsukawa et al. reported that diagnoses of Lisfranc injury on MRI in oblique plane parallel to the ligament with isotropic 3D MRI reliably matched with direct intraoperative observations.^9^ However, MRO is not a dynamic and weight-bearing imaging modality to show functional instability. Another study by Naguib et al, concluded that the accuracy of surgeons’ eye tracking assessment of intraoperative fluoroscopic imaging during stress examination of the tarsometatarsal joint complex in the diagnosis of Lisfranc injuries was reliable.^11^ The Freer elevator test developed by Young et al. has shown promise as a reliable method of intra-operative evaluation of the injured Lisfranc ligament but is limited by possible iatrogenic injury due to the twisting motion and lack of data on its accuracy.^12^ None of these intraoperative methods were either standardized or made reproducible. Our study demonstrated a standardized method using a specific amount of force in a specific direction that can be fundamental in developing devices for intraoperative assessment of the Lisfranc joint, particularly C1-M2 and even C1-C2 as the next step.

A limitation of our study was that we only assessed C1-M2 joint and did not include C1-C2 and other tarsometarsal joints. Moreover, we did not conduct intraoperative measurement and the C1-M2 diastasis to compare with the radiographic measurement. Lastly, the number of our cadavers, though it was based previous studies, was limited, which can lead to reducing the validity and reliability of our measurements while will not affect the purpose of the study. A strong point of our study was the use of different fixation methods that are available in practice, as a means to test our method in different fixation techniques.

## Conclusion

The Listract test is a standardized and reproducible intraoperative examination technique that applies 50N distraction forces in the direction of the C1-M2 ligament. This can help the surgeon assess the joint in a simulated real-life situation under a specific force for distaction and, thus, instability. Future studies with larger cadaver populations to establish cut-off values for both radiographic measurement and intraoperative measurement under Listract test are necessary. In order to use this method intraoperatively, we aimed to design a device that can measure the amount of distraction force, apply 50N, and also measure the amount of diastasis by the surgeon efficiently.

## Data Availability

All data produced in the present study are available upon reasonable request to the authors.

## Acknowledgement

This study was logistically and financially sponsored by FARIL-MGH Center, Ossio Inc., and Depuy Synthes, who helped us provide the instruments and conduct the experiment.

